# Predictors of a 30-day mortality following the first episode of stroke among patients admitted at referral hospitals in Dodoma, central Tanzania: A prospective longitudinal Observational Study

**DOI:** 10.1101/2023.06.20.23291665

**Authors:** Anna M. Chongolo, Alphonce Baraka, Peter M. Mbelele, John Meda, Azan Nyundo

## Abstract

**Background:** Stroke is the leading cause of disability and the second most common cause of death after ischemic heart disease worldwide. A better understanding of the predictors of early post-stroke mortality provides opportunity for interventions that promote favourable post-stroke outcomes.

**Objective:** This study aimed to determine incidence and risk factors associated with 30-day mortality among adult patients admitted with first episode of stroke at referral hospitals in Dodoma.

**Methods:** The study employed a prospective longitudinal observational design. Adult patients with confirmed acute stroke by Computed Tomography scan or Magnetic Resonance Imaging, admitted to Dodoma Referral Hospitals were enrolled in the study. The National Institute of Health Stroke Scale was used to assess stroke severity at baseline. A comparison of risk factors, clinical profiles, and mortality was done using the Chi-square test. A binomial logistic regression model was used to determine the predictors of 30 days mortality in patients with stroke while 30-days probability of survival was estimated using Kaplan-Meier analysis.

**Results:** Out of 226 patients with first-ever stroke, 121(54%) were males and the population mean age was 63(15) years. 140(62%) had Ischaemic stroke subtype, and 154(68%) survived at 30 days of stroke after admission. Patient with history of smoking 2.4 [95% CI (1.0 - 5.6), p = 0.048], loss of consciousness 2.7 [95% CI (1.2 - 6.4; p = 0.019] and unequal pupil size 13.7 [95% CI (4.1 - 58.1, p < 0.001 were significantly more associated with mortality within 30-days. The median survival was 7 (3-9) days, whereas alcohol drinkers and those aged above 60 years had a shorter time to mortality compared to non-alcohol drinkers and those aged < 60 years.

**Conclusion:** The study reveals high incidence of mortality within 30 days after the first episode of stroke, with the highest proportion die within seven days of being hospitalized. Advanced age of 60 years and above, smokers, alcoholic users, and severe stroke at admission warrant special attention. remains the most catastrophic and disabling conditions, with profound residual impairment and a high fatality rate, that puts a significant strain on community health expenditures as well as patients and their families (1–3). Globally, after ischemic heart disease, stroke is the second leading cause of mortality accounting for 11.8% of total deaths (4). Most of stroke related deaths occur in developing countries, accounting for about 87% of stroke deaths (5). Worldwide, one out of six persons will experience a stroke in their lifetime, with 5 to 10% of all stroke victims being under the age of 50 (6). In a 2004-2006 Tanzanian population-based study, the crude stroke incidence was 107.9 per 100,000 for urban and 94.5 per 100,000 for rural areas, and 315.9 and 108.6 per 100,000 for rural and urban respectively following age standardization (7); interestingly, the study highlighted higher incidence of stroke in urban Tanzania compared to developed countries(7).

The most common conventional risk factors for stroke in Africa are hypertension, diabetes, smoking, a sedentary lifestyle, sickle cell disease, African race, an increasing in ageing population and alcohol abuse (8). Meanwhile, over 80% of published studies in Sub-Saharan Africa (SSA) identify hypertension as the most frequently identified risk factor (9). Both the rapid rise of hypertension and the poor control of blood pressure in Africa contribute to an increase in haemorrhagic stroke, which has a worse outcome than ischemic stroke (10,11).

Thirty-day stroke mortality ranges between 3.1 to 9.7 % in high-income countries (12–14); however, it remains higher in Sub-Saharan Africa, ranging between 27 and 46% (7,15–17). Because of a lack of specialized facilities like stroke units, low- and middle-income countries have a greater stroke death rate than industrialized nations(18). Other predictors include premorbid conditions such as diabetes mellitus, advanced age, the severity of stroke on admission, haemorrhagic type of stroke, and infections (5,19,20). Two previous studies done in Tanzania reported a 33.3% and 61.3% in hospital stroke fatality rates, mortality rate was significantly higher in patients with septicaemia, age above 45 years, and aspiration pneumonia(21)

Given the high global prevalence of stroke, there is limited information on the epidemiology, prevention, treatment, and outcome of stroke in African settings and other LMICs (3,22); therefore, limiting the baseline evidence for designing interventions to reduce this burden in developing countries (23). Consequently, the purpose of this study was to determine predictors of early mortality among adult patients admitted with first episode of stroke in referral hospitals in Dodoma.

## Methods

### Study design and population

This was a prospective longitudinal observational study done at the referral hospitals in Dodoma, the capital of Tanzania. Both Dodoma Referral Regional Hospital and Benjamin Mkapa Hospital receive a combined total of 20 and 30 patients with first stroke episode every month. Both are accredited teaching hospitals for the University of Dodoma’s at both undergraduate and residency medical training programs. The Benjamin Mkapa Hospital is equipped with neuroimaging services, such as CT scans and MRIs in case a patient needs a one.

### Sample size and sampling procedure

Using a formula for proportion in a prospective cohort study (24,25) a minimum sample size of 142 was required. A total of 226 samples were collected over nineteen months, from January 2020 to August 2021. Participants who were readily accessible, willing to participate, and met the inclusion criteria were recruited until the required sample size was attained.

### Data collection

Direct interview with patients or guardians to enquire information on socio-demographic, cardiovascular risk factors, predictors of stroke outcomes and medication history. Laboratory tests: while, radiological results were obtained and recorded in the structured questionnaires.

All adult Patients from 18 years who met the inclusion criteria were enrolled after receiving detailed information about the study. Personal information (including sex, age, marital, occupation), past medical history (like hypertension, diabetes mellitus, previous cardiovascular disease and stroke) and lifestyle including alcohol drinking and smoking were collected. Current smoking/alcohol use, defined as those who smoke or take alcohol within the last 12 months from the presentation was collected through direct interviews with the patient and/or immediate guardian (26). The National Institute of Health Stroke Scale(NIHSS) was used to assess severity of stroke based on symptoms and physical examination using specific scores(27); a score of 1-4 was categorized as minor stroke, 5-15 as moderate stroke, 15-20 as moderately severe stroke and 21-42 as severe stroke. Blood Pressure was measured using a brand AD Medical Inc. digital BP machine by placing the cuff on the arm at the level of the heart. Three measurements were taken in 2 minutes apart from the unaffected arm to minimize the effects of changes in tone from the hemiplegic arm(28). Hypertension is defined as SBP ≥140mmHG or DBP ≥90 mmHg or those with a known history of hypertension or on antihypertensive drugs(28,29). An examination of the radial pulse was done on the unaffected limb for the rate and rhythm in one minute to detect the presence of an arrhythmia, a pulse deficit of 10 or more was regarded as atrial fibrillation (30). Waist and hip circumference were measured using a tape measure and recorded in centimetres. Central obesity was defined as waist-hip ratio reached ≥0.90 in men and ≥0.85 in females (31).

Blood glucose was measured using Glucometer Accucheck Active Roche. A diagnosis of DM was made when RBG level of ≥ 11.1mmol/l and confirmed by fasting blood glucose ≥7.0mmol/l or glycosylated haemoglobin≥ 6.5%. Blood samples of about ten millilitres were collected aseptically: 5mls were analysed for Complete Blood Count (CBS) using Automated Haematological analyser CD-RUBY (32) and a total white cell count >11.6,000 was considered as leucocytosis, 5mls analysed for lipid profile using Automated chemistry analyser Erba XL-180(33), and the results were out within 24 hours post admission and total cholesterol of more than 6.2mmol/l (>240mg/dl) termed as hyperlipidaemia. HIV rapid testing was done using the SD Bio line. If the SD Bio line turns positive then a confirmation test using a rapid test Unigold was done.

A 12-lead electrocardiogram (ECG), using the BIONET machine was performed by the principal investigator to look for arrhythmias. Echocardiography (ECHO) was performed by the consultant cardiologist using GE LOGIQC5 Premium and the result was interpreted based on American society of echocardiography guidelines (34).

Non-contrast brain computed tomography using (SOMATOM Definition Flash) or magnetic resonance imaging was performed on all cases with sudden onset of the neurological deficit as a high suspicion of stroke within seven days, patients were transferred to BMH using either a hospital ambulance with the escort nurse and the results were interpreted by a consultant radiologist (35). Stroke outcome was assessed using a modified Rankin Scale(36) at 24 hours, 7 days, 14 days and 30 days from admission.

### Data analysis

The study population characteristics were summarized as mean and standard deviation, median and inter-quartile range for continuous variables while categorical variables were summarized as frequencies and proportions. Chi-square test was used to assess baseline characteristics of stroke patients and the association with 30-days mortality. Binary logistic regression was used to compute for the predictors of 30-days mortality, only variables that met a 20% statistical significance at univariate level (Hosmer, Lemeshow, & Sturdivant, 2013) were selected for multivariable Logistic regression analysis to determine independent predictors for 30 – days mortality summarised as adjusted odds ratio (aOR), 95% confidence interval (CI) and statistical significance set at 5%(α=0.05) level. Time to mortality by associated factors using log rank test, and statistical significance was set at a p value less than 0.05.

### Ethical considerations

The ethical clearance was provided by the University of Dodoma Institutional Review Board under the Directorate of Research and Publications (reference number MA.84/261/02). Following that, the administrative departments of Benjamin Mkapa and Dodoma Regional Referral Hospitals approved data collection with reference numbers AC.83/119/01/89 and PB.22/130/02/04, respectively. Participants or next of kin were informed that their participation was fully optional and that they may opt out at any moment without interfering with the standard routine care. Participants were asked to provide a written or verbal informed consent under the witness of the close relative. For those who could not provide informed consent, a custodian who had to be a close family member or guardian provided the assent on behalf.

### Socio-demographic and clinical characteristics of participants

Of all 242 patients presented with stroke, 226 (93.4%) met the criteria for first ever stroke and able to provide informed consent, refer figure 1 for the reasons of excluding 16. Of these 226 patients, 121 (54%) were male, the mean (SD) age was 63 (15) years and the median NIHSS (IQR) was 14 (11–20). A total of 140 (62%) patients had an ischemic stroke, 54(24%) were cigarette smokers, 189 (84%) had hypertension, 38 (17%) had diabetes, just 7 (3%) patients were living with HIV/AIDS and on regular use of medications (Table 1).

**Figure 1.**
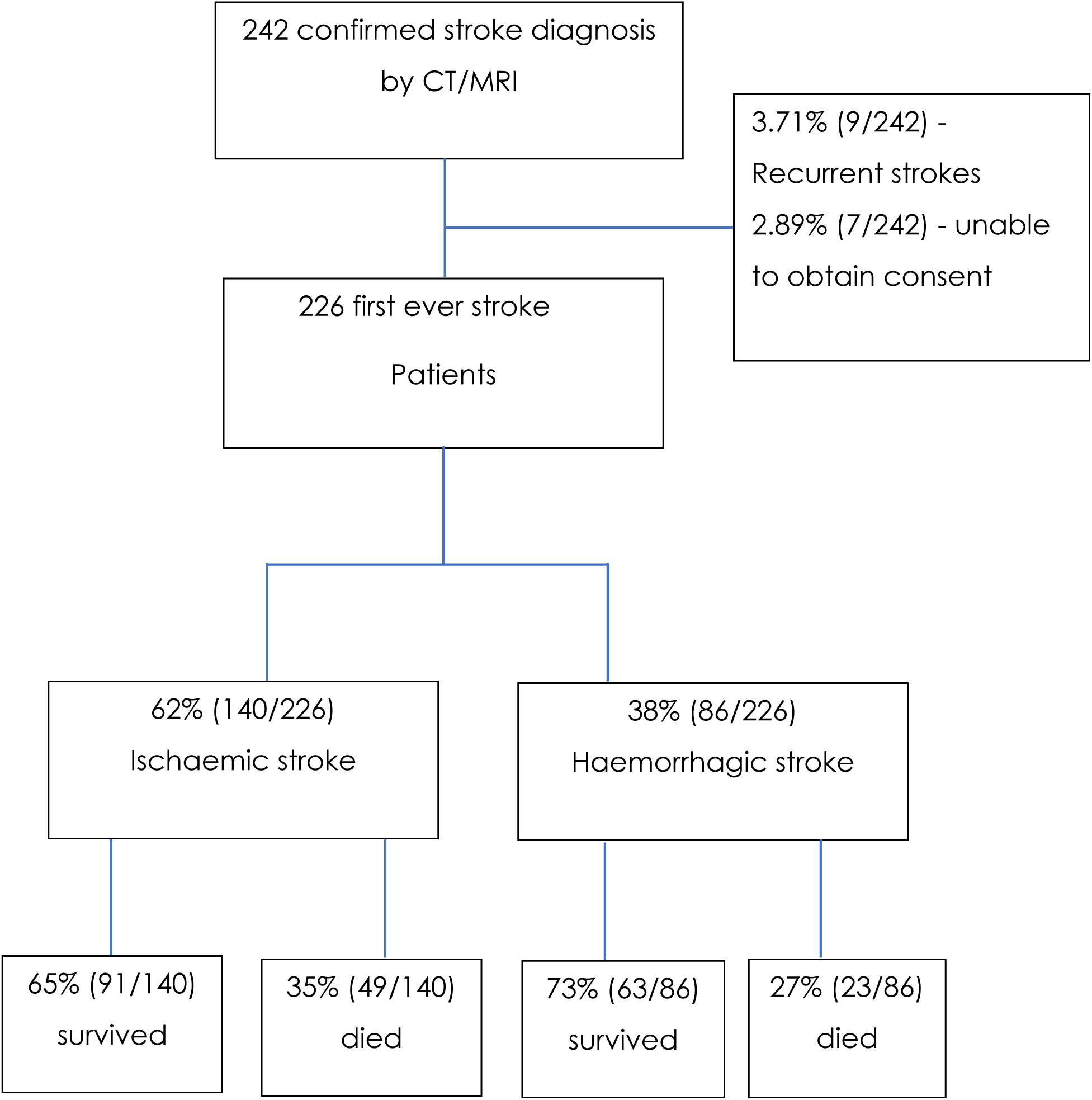
Recruitment and clinical outcome of patients with stroke.

**Table 1.** Baseline characteristics of patients who died versus survived due to stroke.

| Table 2 Baseline characteristics of patients who died versus survived due to stroke |  |  |  |  |  |
| --- | --- | --- | --- | --- | --- |
| Variables | Category | All<br>(N = 226) | Survived<br>(n = 154) | Died<br>(72) | P-value |
| Gender, n (%) | Female | 105 (46) | 71 (46) | 34 (47) | 0.989 |
|  | Male | 121 (54) | 83 (54) | 38 (53) |  |
| mean age,<br>(SD) | mean | 63 (15) | 61 (13) | 69 (96) | < 0.001 |
| HIV* | Positive | 7 | 6 | 1 | 0.425 |
|  | Negative | 137 | 90 | 47 |  |
| Marriage* | Single | 25 | 14 | 11 | 0.312 |
|  | Married | 119 | 82 | 37 |  |
| Education | No formal | 48(21) | 24(16) | 24(33) | 0.015 |
|  | Primary | 107(47) | 77(50) | 30(42) |  |
|  | Secondary | 38(17) | 26(17) | 12(17) |  |
|  | Tertiary | 33(15) | 27(17) | 6(8) |  |
| Occupation* | Dependent | 23 | 10 | 13 | 0.009 |
|  | Employee | 22 | 19 | 3 |  |
|  | Peasant | 99 | 67 | 32 |  |
| Smoking | Yes | 54 (24) | 31 (20) | 23(32) | 0.076 |
|  | No | 172 (76) | 123 (80) | 49(68) |  |
| Alcohol drink | Yes | 91(40) | 58(38) | 33 (46) | 0.307 |
|  | No | 135(60) | 96(62) | 39 (54) |  |
| Hypertension | Yes | 189 (84) | 126(82) | 63(88) | 0.377 |
|  | No | 37 (16) | 28(18) | 9(12) |  |
| Diabetes | Yes | 38 (17) | 25(16) | 13(18) | 0.881 |
|  | No | 188(83) | 129(84) | 59(82) |  |
| Hyperglycemia | Yes | 52(23) | 31(20) | 21(29) | 0.182 |
|  | No | 174(77) | 123(80) | 51(71) |  |
| Aspiration<br>Pneumonia | Yes | 62 (27) | 42 | 20 | < 0.001 |
|  | No | 164 (73) | 134 | 30 |  |
| Stroke Type per<br>CT | Haemorrhagic | 86 (38) | 63(41) | 23(32) | 0.252 |
|  | Ischemic | 140 (62) | 91(59) | 49(68) |  |
| Headache | Yes | 124(55) | 81(53) | 43(60) | 0.249 |
|  | No | 102(45) | 73(47) | 29(40) |  |
| Aphasia | Yes | 142 (63%) | 76(49) | 66(92) | < 0.001 |
|  | No | 84 (37%) | 78(51) | 6(8) |  |
| Vomiting | Yes | 48(21) | 29(19) | 19(26) | 0.263 |
|  | No | 178(79) | 125(81) | 53(74) |  |
| Loss of consciousness | Yes | 87(38) | 35(23) | 52(72) | < 0.001 |
|  | No | 139(62) | 119(77) | 20(28) |  |
| Median NIHSS (IQR) | Median | 14 (11 – 20) | 13 (9 – 17) | 19 (12 – 23) | < 0.001 |
| Focal neurologic deficit | Yes | 222(98) | 151(98) | 71(99) | 0.997 |
|  | No | 4(2) | 3(2) | 1(1) |  |
| Dysarthria | Yes | 73(32) | 69(45) | 4(6) | < 0.001 |
|  | No | 153(68) | 85(55) | 68(94) |  |
| Seizure | Yes | 48(21) | 24(16) | 24(33) | 0.004 |
|  | No | 178(79) | 130(84) | 48(67) |  |
| Pupils examination | Normal | 200(89) | 149(97) | 51(71) | <0.001 |
|  | Anisocoria | 14(6) | 3(2) | 11(15) |  |
|  | Pinpoint | 12(5) | 2(1) | 10(14) |  |
| median RBG (IQR) |  | 6.9 (6.1 – 8.9) | 6.8 (6.0 – 8.1) | 7.5 (6.4 – 10.1) | 0.011 |
| Mean/medial Cholesterol |  | 5.2 (4.5– 6.5) | 5.2 (4.3 – 6.5) | 5.4 (4.7 – 6.3) | 0.338 |
| Mean/medial LDH |  | 2.2 (1.8 – 2.9) | 2.2 (1.8 – 2.9) | 2.3 (1.9 – 3.1) | 0.116 |
| Mean/medial Triglyceride |  | 1.5 (1.1 – 2.1) | 1.4 (1.1 – 2.1) | 1.7 (1.2 – 2.1) | 0.133 |

### Predictors of 30-day mortality after first episode of stroke

The 30-day mortality rate among 226 patients with their first episode of stroke was 72 (32%). Under adjusted logistic regression model, cigarette smokers AOR:3.2 [95% CI (1.3 – 7.9), p = 0.009], those presented with loss of consciousness AOR:2.7 [95% CI (1.2 - 6.4); p = 0.019], abnormal pupil size AOR:12.1 [95% CI (3.4 – 53.6), p < 0.001] and aspiration pneumonia AOR:5.2 [95% CI (2.2 – 12.6); p = 0.001] had significantly higher odds of mortality at thirty days post stroke, (Table 2).

**Table 2:**
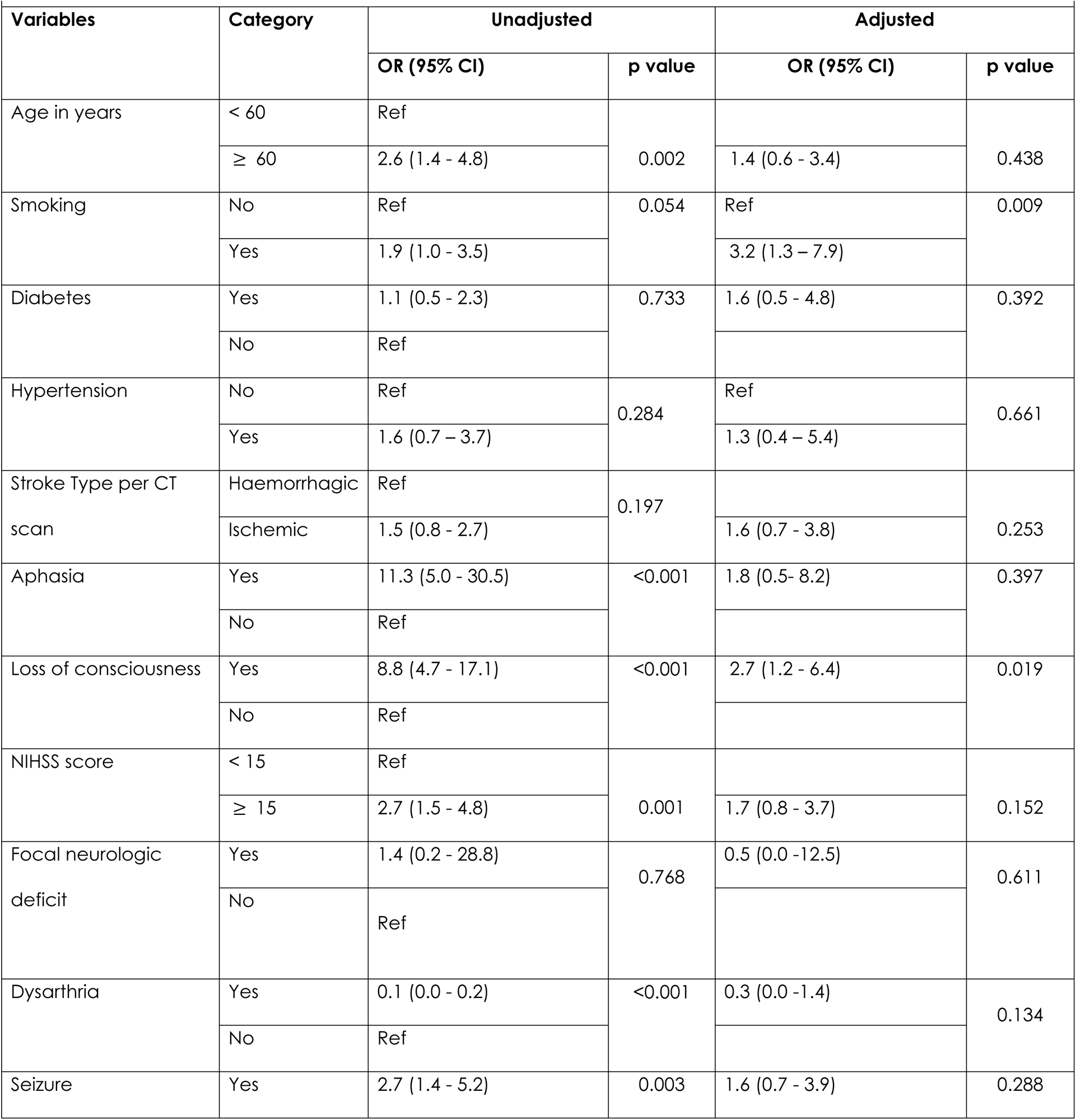
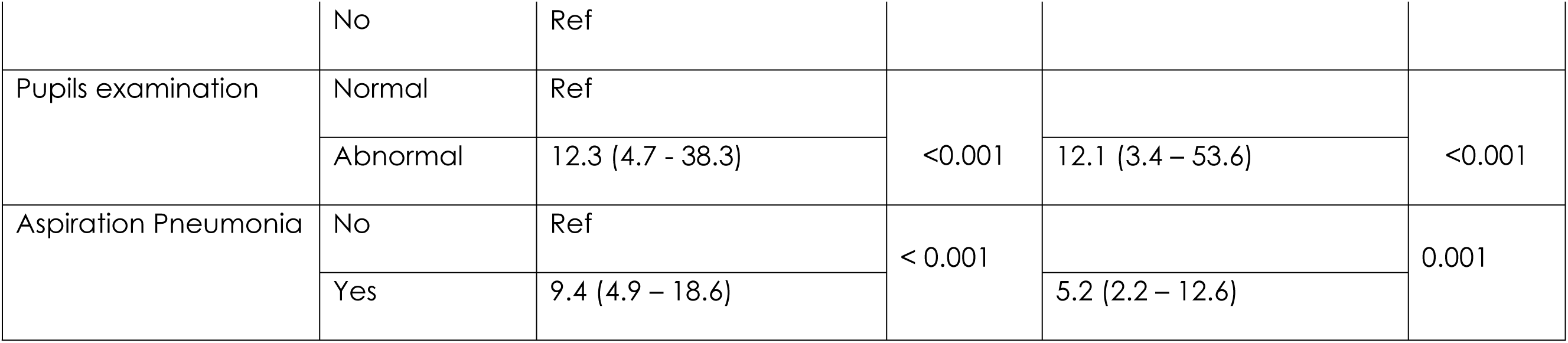
Predictors of 30 days mortality in patients with stroke.

### Time to mortality by clinical patterns and associated risks

The overall mortality in 226 patients presenting with the first episode of stroke was 72 (32%). Their median (IQR) time to mortality was 7 (3–9) days. Whereas alcohol drinkers and those aged above 60 years had a shorter time to mortality compared to non-alcohol drinkers and those aged < 60 years (Figure 2D & 2F, p ≤ 0.041). There were no significant differences in time to mortality among those with or without hypertension, smoking, ischemic and haemorrhagic stroke (Figure 2A – 2C, p > 0.05). The details of time to stroke-related mortality among patients by risk factors and clinical presentations on admission are in Figures 2(A-G) and 3.

**Figure 2.**
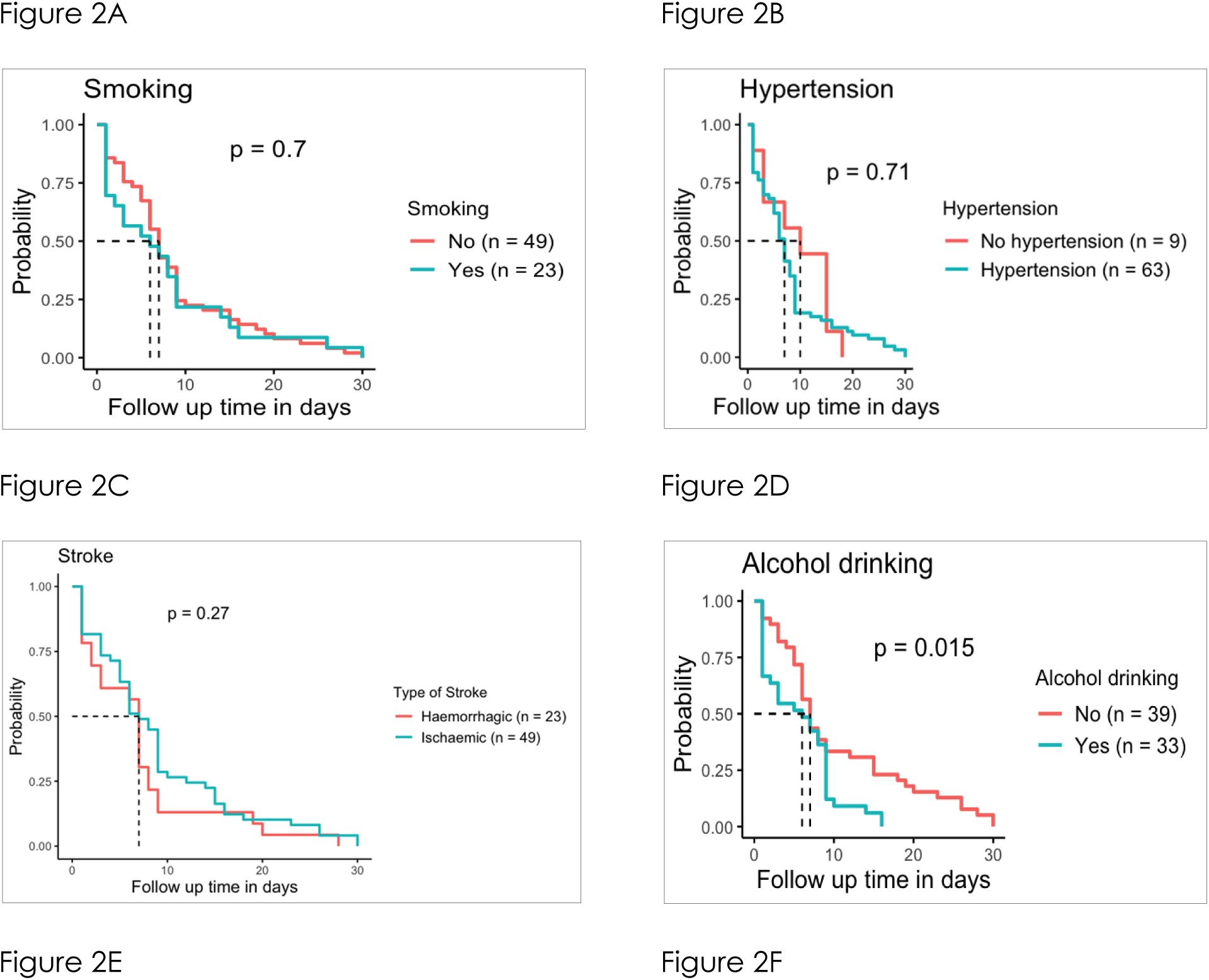
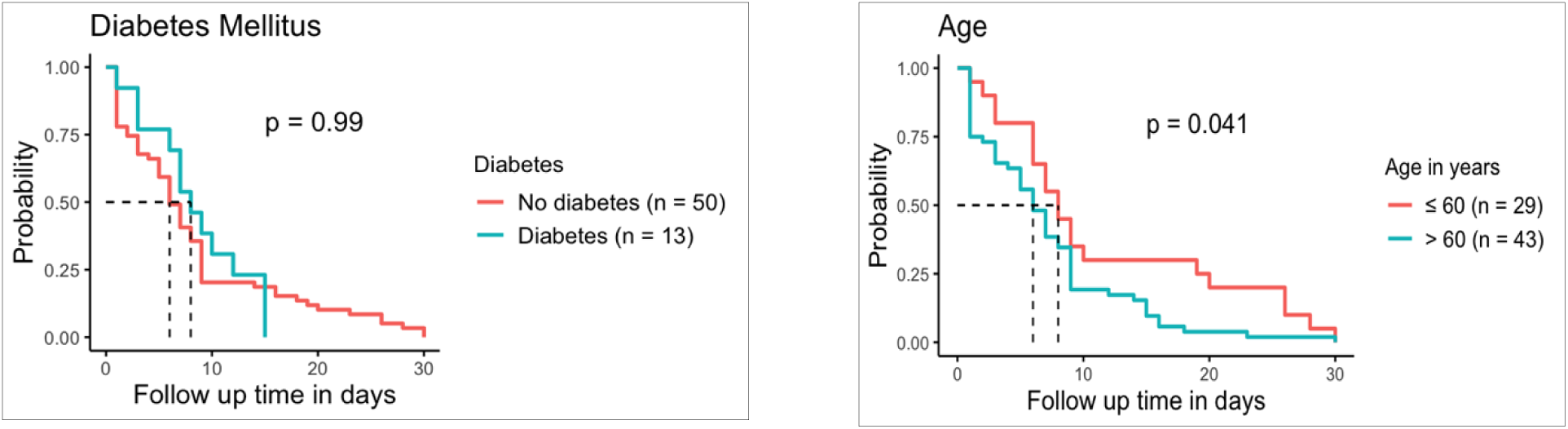
Baseline variables for time to stroke-related death in patients at risk (n = 72)

**Figure 3 (A-G).**
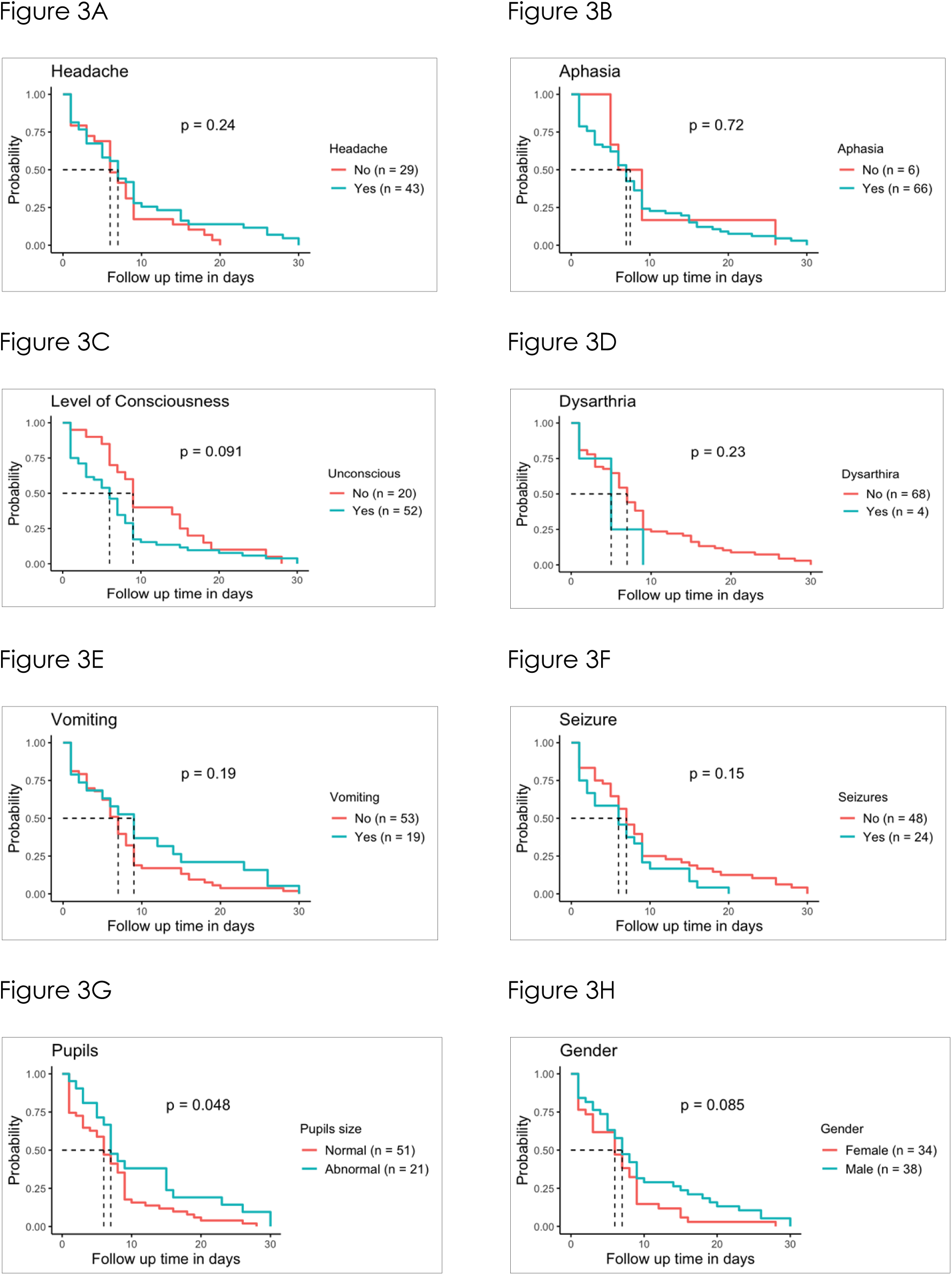
Baseline factors associated to time to stroke-related mortality among patients at risk (n = 72)

## Discussion

This study found ischemic stroke is a more prevalent type (62%), this is similar to a ratio of 60:40 observed at Muhimbili National hospital in Tanzania for ischemic stroke and haemorrhagic stroke, respectively (10,37). Considering variability in ratio of ischemic to haemorrhagic stroke, ischemic stroke remains prevalent in developed and underdeveloped countries (11,38). While the higher risk of intracerebral haemorrhage in low-income and upper middle-income nations may be attributable to the high risk of uncontrolled hypertension in these countries (39), our study also shows 84% of participants were hypertensive with poor blood pressure control. This is consistent with previous findings showing that 52% of Americans (a developed country) have their hypertension under control, compared to 5% to 10% in African population(40). The lack of awareness, access to care, and adherence to implementable hypertension management recommendations in low and middle income countries may be a major contributor to this difference (41).

The 30-day mortality rate in this study was 32%, which was comparable to previous reports from Uganda(42), and Ghana(43), where the mortality rates were 43.8%, and 43%, respectively. The relatively higher mortality rate (61.3%) in previous study conducted in Dar es salaam-Tanzania(37) might be attributed to the inclusion of more than half of participants with severe stroke (median NIHSS was 19) despite the mean age of the study populations was nearly identical to our study population. Our findings also showed the 30-day mortality rate to be 15% higher than in industrialized countries (44–46), owing to a lack of knowledge about stroke signs and symptoms resulting into late hospital presentation(47), about which the mean duration the patient presented to the hospital since the onset of stroke was 3 days in a previous study done in Tanzania (48) similar to the observed mean duration for this study. Furthermore, the low and middle income countries have poor neurosurgical outcome of stroke, with few studies reporting up to 55% mortality rate (49) owing to lack of access to advanced neurosurgical procedures compared to developed nations.

We found that current smoking status, loss of consciousness, unequal pupil size at presentation and post stroke aspiration pneumonia were independent determinants of stroke mortality at 30 days. The impact of smoking on stroke outcomes is consistently reported (50) in diverse settings including the United states and China (51–53); however, the association between smoking and stroke-related mortality is not always delineated given the disparity in access to advanced stroke units that offer standard and timely stroke management in the acute setting in developed countries. For example, unlike the resource limited settings, thrombolytic agents are generally provided which may improve their stroke severity score at baseline and hence improve outcome are generally provided in the advanced units of developed world (54). Given that, smokers tend to have higher NIHSS score at admission (55), are more likely to have hypertension, abdominal obesity, dyslipidemia, atrial fibrillation, and diabetes (56), the difference in improvement may particularly be observed among smokers who receive thrombolytic agents compared to those who do not. it is hypothesized that smoking reduces the release of endogenous tissue-type plasminogen activator, leading to an increase in fibrinogen levels and an increase in fibrin-rich thrombi, thereby increasing susceptibility to exogenous tissue-type plasminogen activator therapy (57). Furthermore, the risk of vascular events such as heart attacks and strokes appear to be amplified when these risk factors coexist (58). Loss of consciousness is a good proxy for the severity of a stroke, and both have bee n associated with an increased risk of stroke mortality in the early stages (59). In a previous study done in the United States, it was shown that less severe types of stroke were linked to an 80% drop in stroke-related deaths, which supports the findings elsewhere (60). We found that stroke patient with loss of consciousness at presentation were three times higher odds of dying within 30 days. Similar findings were reported in previous study conducted in Nigeria; patient with Glasgow coma scale < 10 and with NIHSS ≥ 16 were more likely to die within 30 days and other studies in Africa and western countries also show severity of stroke predict high mortality rate (61,62).

As observed previous, unequal pupil size/anisocoria on admission predicted death within 30 days compared those with normal pupil (59,63). Unequal pupil size is common presentation of stroke in vertebral-basilar circulation mostly due to expanding aneurysm of posterior communicating artery that compress externally located pupil fibres of oculomotor nerve (64). With the mortality of 85%, vertebral basilar-stroke carries higher risk of mortality compared to stroke in anterior circulation; few things may explain this phenomena, vertebral-basilar stroke presents with symptoms that resembles non-strokes symptoms such as vertigo or nausea and thus delaying timely-dependent interventions[63,64]. Given, its involvement of the brain stem and cerebellum, vestibular-cerebellar strokes are also associated with severe disabilities including hemiplegia or quadriplegia, dysarthria, dysphagia, gaze abnormalities and cranial neuropathies secondary due to multisystem involvement[64]. Although, variables such as higher presenting NIHSS typically present which typically associate with poorer functional outcome are more common in anterior circulation stroke than in posterior circulation stroke(67), the NIHSS detects posterior circulation deficits with less sensitivity than anterior circulation deficits(68), therefore, its utility in predicting poor outcome posterior circulation stroke is rather limited.

We found that post stroke aspiration pneumonia was common (27%), and was an independent predictor of early stroke mortality. Our findings are consistent with previous studies conducted in Sub-Saharan Africa; published evidence from this region indicates that its incidence ranges from 13% to 34% and that it may be responsible for up to 45% of inpatient stroke-related mortality (21,69,70). In the United States, post-stroke aspiration pneumonia accounts for up to 13% of cases and up to 10% of deaths resulting from strokes (71). Stroke patients frequently develop aspiration pneumonia due to dysphagia and decreased mental status (71). Post stroke aspiration pneumonia lengthens hospital stays, induces fevers and systemic inflammation, and can develop into sepsis and acute respiratory failure (72,73). Inpatient stroke centres in the United States and other high-resourced settings use protocols to reduce the risk of post stroke aspiration pneumonia, such as early swallow screening, nil per os (NPO), and Nasogastric tube (NGT) feeding in patients with prohibitive dysphagia, but resource-limited settings often lack adequate human resource for close nursing care and swallowing therapists to ensure safe nutrition after an acute neurologic insult that impairs mental status or oropharyngeal coordination (74,75). In sub-Saharan Africa, limited access to NGTs, parenteral feeding formulae, food texture change, difficulty to elevate the (head-of-bed) HOB in many hospitals, and vigorous oral feeding by bedside family caregivers may increase risk of aspiration.

Majority of hospitalized stroke patient died within 7 days which was found to be comparable with 7 days in a study done at Debre Markos Comprehensive Specialized Hospital-Northwest Ethiopia(76), 4.5 days at Hawassa University Referral Hospital-South Ethiopia(77), and 4 days at Ayder Comprehensive Specialized Hospital-Northern Ethiopia(78). Early stroke deaths that occur within the first week of stroke onset was primarily influenced by neurological dysfunction (79), initial stroke severity and early neurological deterioration (20).

The study had a number of limitations. First, this was a hospital-based study, which may not accurately reflect the reality given the lack of access to hospital for many critically ill patients who might have died before reaching hospital. In addition, the hospital-based study is susceptible to referral bias because the majority of acute stroke patients that visit our hospital are from the health facilities proximal to the hospitals. Thus, referral biases and the method of convenience sampling may not adequately be generalizable to the community.

## Conclusion

In central Tanzania, the 30-day rate of mortality after stroke is high, and most patients die within seven days of being hospitalized. Stroke patients over the age of 60, smokers, alcoholic drinkers, and patients who arrive at the hospital with loss of consciousness and variable pupil size should be given special attention.

## Data Availability

Data will be shared upon request

## Acknowledgements

We thank all the study participants and internal medicine team from DRRH and BMH for the support during data collection.

## Author contributions

Conceptualized and designed of the study: AC, JM and AN.

Data collection and analysis: AC, PM and AN

Report writing: AC, PM, AN, JM.

Manuscript writing: AC, PM, AB, and AN, ECHO done by: AC and JM

